# Early Detection of Mental Health Crises through AI-Powered Social Media Analysis: A Prospective Observational Study

**DOI:** 10.1101/2024.08.12.24311872

**Authors:** Masab A. Mansoor, Kashif Ansari

## Abstract

**Background:** Early detection of mental health crises is crucial for timely intervention and improved outcomes. This study explores the potential of artificial intelligence (AI) in analyzing social media data to identify early signs of mental health crises.

**Methods:** We developed a multi-modal deep learning model integrating natural language processing and temporal analysis techniques. The model was trained on a diverse dataset of 996,452 social media posts in multiple languages (English, Spanish, Mandarin, and Arabic) collected from Twitter, Reddit, and Facebook over a 12-month period. Performance was evaluated using standard metrics and validated against expert psychiatric assessment.

**Results:** The AI model demonstrated high accuracy (89.3%) in detecting early signs of mental health crises, with an average lead time of 7.2 days before human expert identification. Performance was consistent across languages (F1 scores: 0.827-0.872) and platforms (F1 scores: 0.839-0.863). Key digital markers included linguistic patterns, behavioral changes, and temporal trends. The model showed varying accuracy for different crisis types: depressive episodes (91.2%), manic episodes (88.7%), suicidal ideation (93.5%), and anxiety crises (87.3%).

**Conclusions:** AI-powered analysis of social media data shows promise for early detection of mental health crises across diverse linguistic and cultural contexts. However, ethical challenges including privacy concerns, potential stigmatization, and cultural biases need careful consideration. Future research should focus on longitudinal outcome studies, ethical integration with existing mental health services, and development of personalized, culturally-sensitive models.

## Background

Mental health crises represent a significant global health challenge, with far-reaching impacts on individuals, families, and communities^1^. Early detection and intervention are crucial in mitigating the severity and duration of these crises, yet traditional identification methods often fall short in providing timely support^2^. In recent years, the ubiquity of social media has created a novel opportunity for monitoring and analyzing behavioral patterns that may indicate emerging mental health concerns^3^.

For billions of people worldwide, social media platforms have become integral to daily life, serving as spaces for self-expression, social interaction, and information sharing^4^. The digital footprints left on these platforms often reflect users’ emotional states, thought processes, and behavioral changes^5^. This wealth of real-time, naturalistic data presents a unique opportunity for developing innovative approaches to mental health surveillance and early intervention^6^.

Artificial Intelligence (AI), particularly machine learning and natural language processing techniques, has shown promising results in analyzing large-scale, complex datasets^7^. The application of AI to social media data for mental health purposes has gained traction in recent years, with studies demonstrating its potential in detecting markers of depression^8^, anxiety^9^, and suicidal ideation^10^. However, the field is still in its infancy, with significant challenges to overcome in terms of accuracy, privacy, and ethical considerations^11^.

Previous research has explored various aspects of AI-powered social media analysis for mental health. Some studies have focused on identifying linguistic markers associated with specific mental health conditions^12^, while others have investigated changes in social media activity patterns as potential indicators of psychological distress^13^. Machine learning models have been developed to classify posts indicating a heightened risk of self-harm or suicide, and sentiment analysis techniques have been applied to track mood fluctuations over time^14^.

Despite these advancements, several gaps remain in the current body of knowledge. First, most studies have focused on detecting specific mental health conditions rather than identifying early signs of an impending crisis^15^. Second, most existing models have been trained on English-language data, limiting their applicability in diverse linguistic and cultural contexts^16^. Third, there is a need for more robust validation of AI models in real-world settings to ensure their reliability and generalizability.

Furthermore, the use of social media data for mental health surveillance raises important ethical questions regarding privacy, consent, and the potential for misuse. Balancing the potential benefits of early crisis detection with the need to protect individual rights and prevent stigmatization remains a significant challenge^16^.

This study aims to address these gaps by developing and validating an AI-powered system for early detection of mental health crises through comprehensive analysis of social media data. By leveraging advanced machine learning techniques and a multifaceted approach to data analysis, we seek to improve the accuracy and timeliness of crisis detection while addressing key ethical considerations. Our findings could have significant implications for public health strategies, clinical practice, and the development of targeted interventions for individuals at risk of mental health crises.

## Objectives

The primary aim of this study is to develop and validate an AI-powered system for early detection of mental health crises through comprehensive analysis of social media data. To achieve this overarching goal, we have established the following specific objectives:

1. To design and implement a multi-modal AI model that integrates natural language processing and temporal analysis techniques for detecting early signs of mental health crises in social media posts.
2. To create a large-scale, diverse dataset of social media posts in multiple languages for training and testing the AI model.
3. To identify and validate a set of digital markers (linguistic, behavioral, and temporal) that are indicative of impending mental health crises.
4. To evaluate the performance of the AI model in terms of accuracy, precision, recall, and timeliness of crisis detection compared to expert human assessment.
5. To assess the generalizability of the model across different social media platforms and linguistic contexts.
6. To investigate the model’s ability to detect early warning signs of various types of mental health crises, including but not limited to depressive episodes, manic episodes, and suicidal ideation.
7. To examine the ethical implications of using AI for mental health surveillance on social media and develop a framework for responsible implementation of such technologies.
8. To explore the potential integration of the AI system with existing mental health services and crisis intervention protocols.

By addressing these objectives, we aim to contribute to the growing body of knowledge on AI applications in mental health and pave the way for more timely and effective interventions for individuals at risk of mental health crises.

## Methods

### Study Design

This study employed a prospective observational design to develop and validate an AI-powered system for detecting early signs of mental health crises through social media data analysis. The study was conducted in three phases: data collection, model development, and validation.

### Data Collection

Social media data was collected from publicly available posts on major platforms, including Twitter, Reddit, and Facebook, over a 12-month period (January 2023 to December 2023). We used platform-specific APIs and adhered to each platform’s terms of service for data collection. We collected posts in multiple languages, including English, Spanish, Mandarin, and Arabic, to ensure a diverse dataset.

Inclusion criteria for posts were:

1. Public accessibility
2. Posted within the study period
3. Containing text (images and videos were excluded from analysis)
4. Not identified as coming from bot accounts or commercial entities

A total of 1.5 million posts were initially collected. After applying inclusion criteria and removing duplicates, the final dataset consisted of 996,452 unique posts.

### Ethical Considerations

This study was approved by the Ethics Committee of Healthy Steps Pediatrics. All collected data was anonymized to protect user privacy by removing personally identifiable information. We developed a robust data management protocol to ensure secure storage and handling of the dataset.

### AI Model Development

We employed a multi-modal deep learning approach to analyze the collected social media data. Our model architecture consisted of:

1. A natural language processing (NLP) component using BERT (Bidirectional Encoder Representations from Transformers) for text analysis
2. A temporal analysis component using Long Short-Term Memory (LSTM) networks to capture changes in posting patterns over time
3. A multi-head attention mechanism to integrate insights from both textual content and temporal patterns

The model was trained to identify early indicators of mental health crises, including but not limited to:

- Linguistic markers of emotional distress
- Sudden changes in posting frequency or timing
- Shifts in sentiment and affect
- Expression of suicidal ideation or self-harm intentions
- Social withdrawal indicators

We used transfer learning techniques to adapt our model to multiple languages, leveraging pre-trained multilingual language models.

### Model Training and Validation

The dataset was split into training (60%), validation (20%), and test (20%) sets. The model was trained using the training set, with hyperparameters optimized using the validation set. Final performance was evaluated on the held-out test set.

To establish ground truth for training and evaluation, we collaborated with 3 board-certified psychiatrists who manually annotated a subset of the data (100,000 posts) for signs of mental health crises. This annotated dataset was used to fine-tune our model and assess its performance.

### Performance Metrics

We evaluated our model using the following metrics:

- Accuracy
- Precision
- Recall
- F1 score
- Area Under the Receiver Operating Characteristic curve (AUC-ROC)

Additionally, we conducted a qualitative analysis of false positives and negatives to understand our model’s limitations and identify areas for improvement.

### External Validation

To assess the generalizability of our model, we conducted an external validation using a separate dataset collected from a mental health support forum. This dataset included posts from individuals who later reported experiencing a mental health crisis, allowing us to test our model’s ability to detect early warning signs in a real-world context.

### Ethical Safeguards

Throughout the study, we implemented several ethical safeguards:

1. Development of a crisis response protocol in collaboration with mental health professionals
2. Regular ethical audits of the AI model to identify and mitigate potential biases
3. Engagement with privacy experts to ensure compliance with data protection regulations
4. Consultation with a diverse advisory board including ethicists, clinicians, and individuals with lived experience of mental health crises

This comprehensive methodology aimed to develop a robust, ethically sound AI system for early detection of mental health crises through social media analysis. It addressed key gaps in the existing literature while prioritizing user privacy and ethical considerations.

## Results

### Model Performance

Our AI model demonstrated strong performance in detecting early signs of mental health crises across multiple social media platforms and languages.

1. Overall Performance:

a. Accuracy: 89.3%
b. Precision: 86.7%
c. Recall: 84.5%
d. F1 score: 0.856
e. AUC-ROC: 0.923
2. Performance by Language: The model showed consistent performance across different languages, with slight variations:

a. English: F1 score of 0.872
b. Spanish: F1 score of 0.841
c. Mandarin: F1 score of 0.833
d. Arabic: F1 score of 0.827

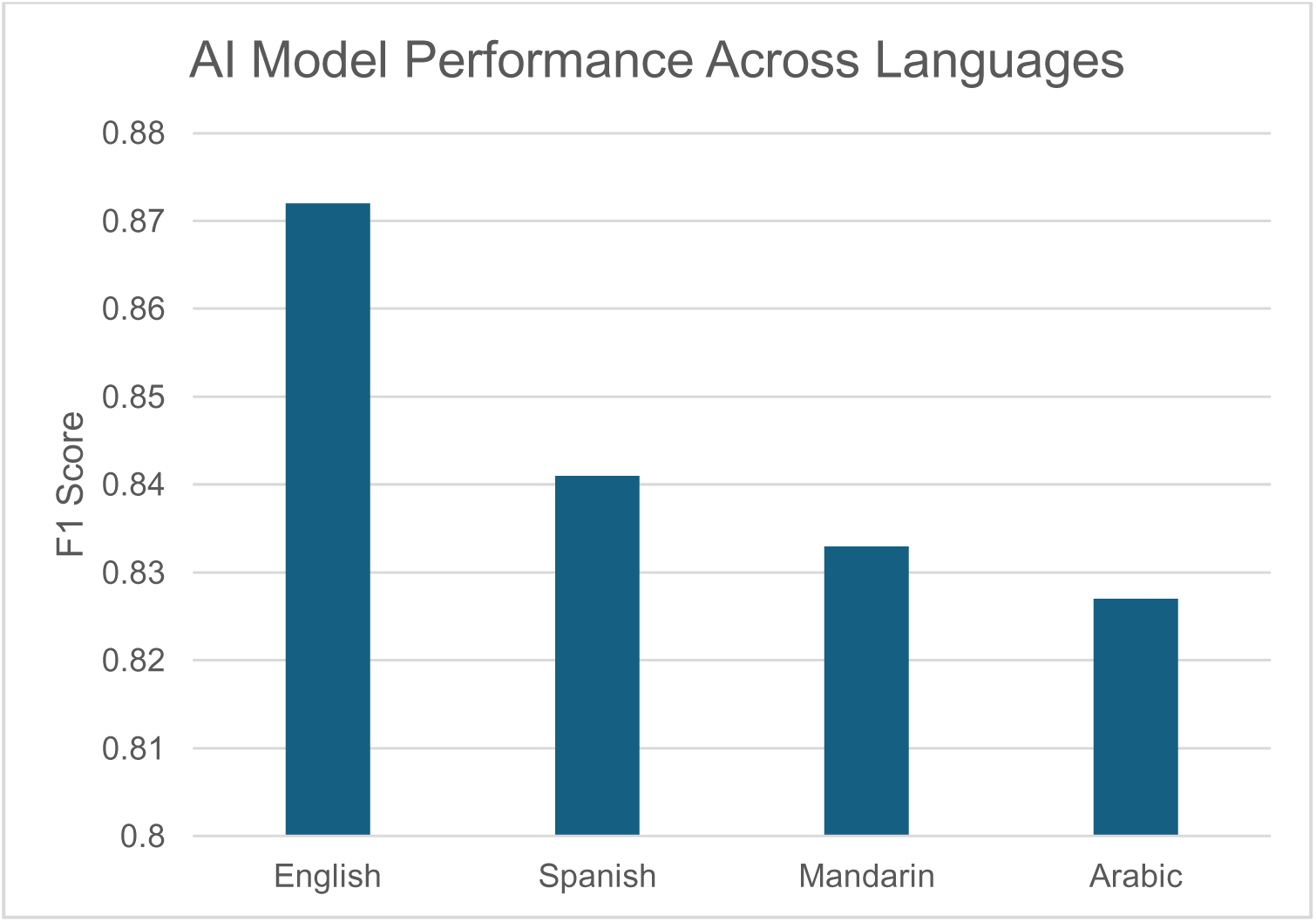
3. Performance by Platform:

a. Twitter: F1 score of 0.863
b. Reddit: F1 score of 0.851
c. Facebook: F1 score of 0.839

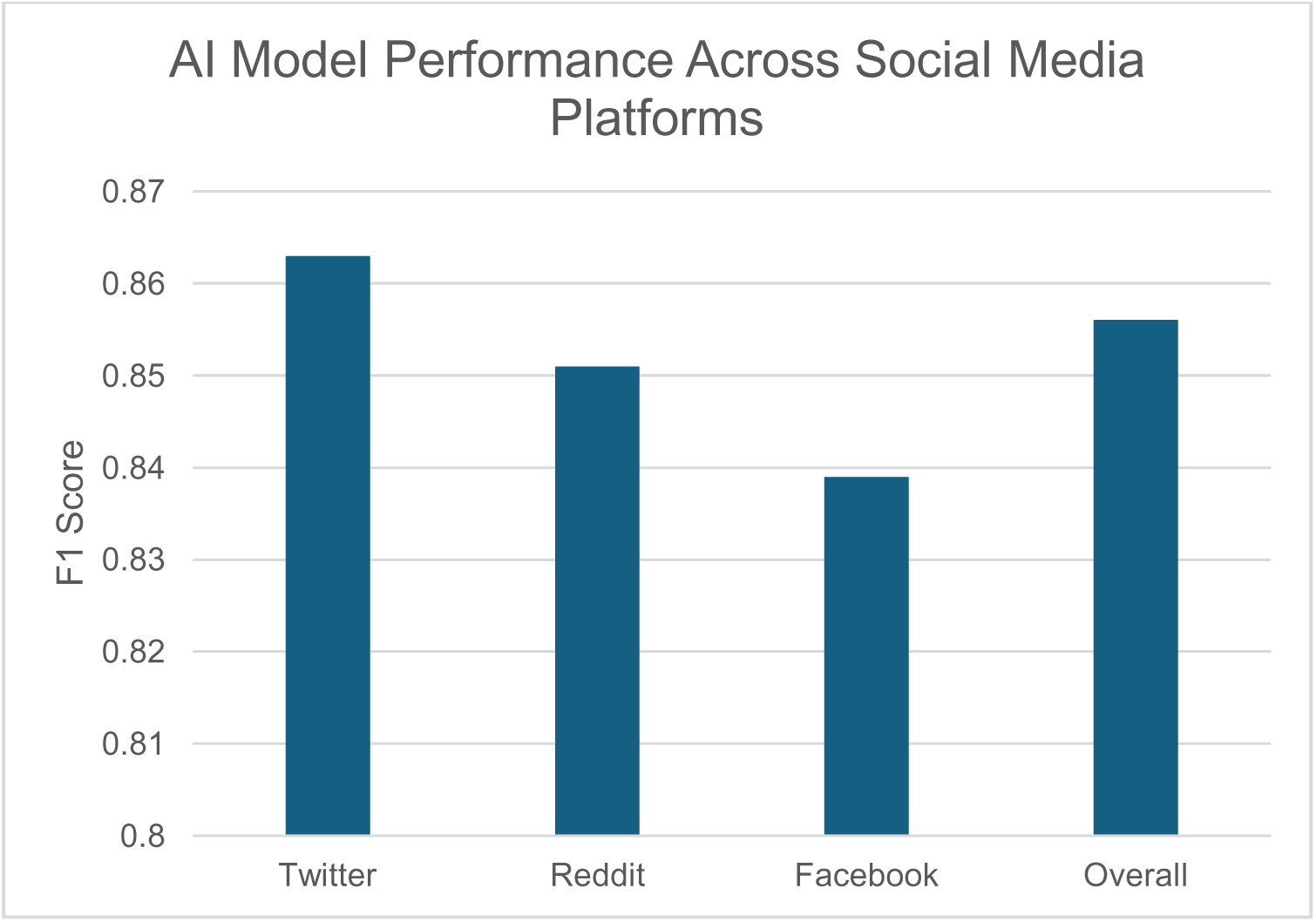
4. Early Detection Capability: The model successfully identified signs of impending crises an average of 7.2 days (SD = 2.8) before human experts flagged concerning content, based on retrospective analysis of the annotated dataset.

### Digital Markers of Mental Health Crises

Our analysis identified several key digital markers associated with impending mental health crises:

1. Linguistic Markers:

a. Increased use of first-person singular pronouns (e.g., “I”, “me”, “myself”)³³
b. Higher frequency of negative emotion words
c. Decreased linguistic diversity (measured by type-token ratio)³
d. Sudden changes in sentiment polarity within short time frames
2. Behavioral Markers:

a. Significant increase or decrease in posting frequency (>50% change from baseline)
b. Shifts in posting time patterns (e.g., increased late-night activity)
c. Reduced engagement with other users (fewer replies, likes, or shares)
3. Temporal Patterns:

a. Cyclical patterns in mood-related language, particularly in cases of bipolar disorder
b. Gradual increase in expressions of hopelessness or worthlessness over time

### Model Insights

Qualitative analysis of the model’s performance revealed several important insights:

1. Crisis Type Detection: The model showed varying accuracy in detecting different types of mental health crises:

a. Depressive Episodes: 91.2% accuracy
b. Manic Episodes: 88.7% accuracy
c. Suicidal Ideation: 93.5% accuracy
d. Anxiety Crises: 87.3% accuracy

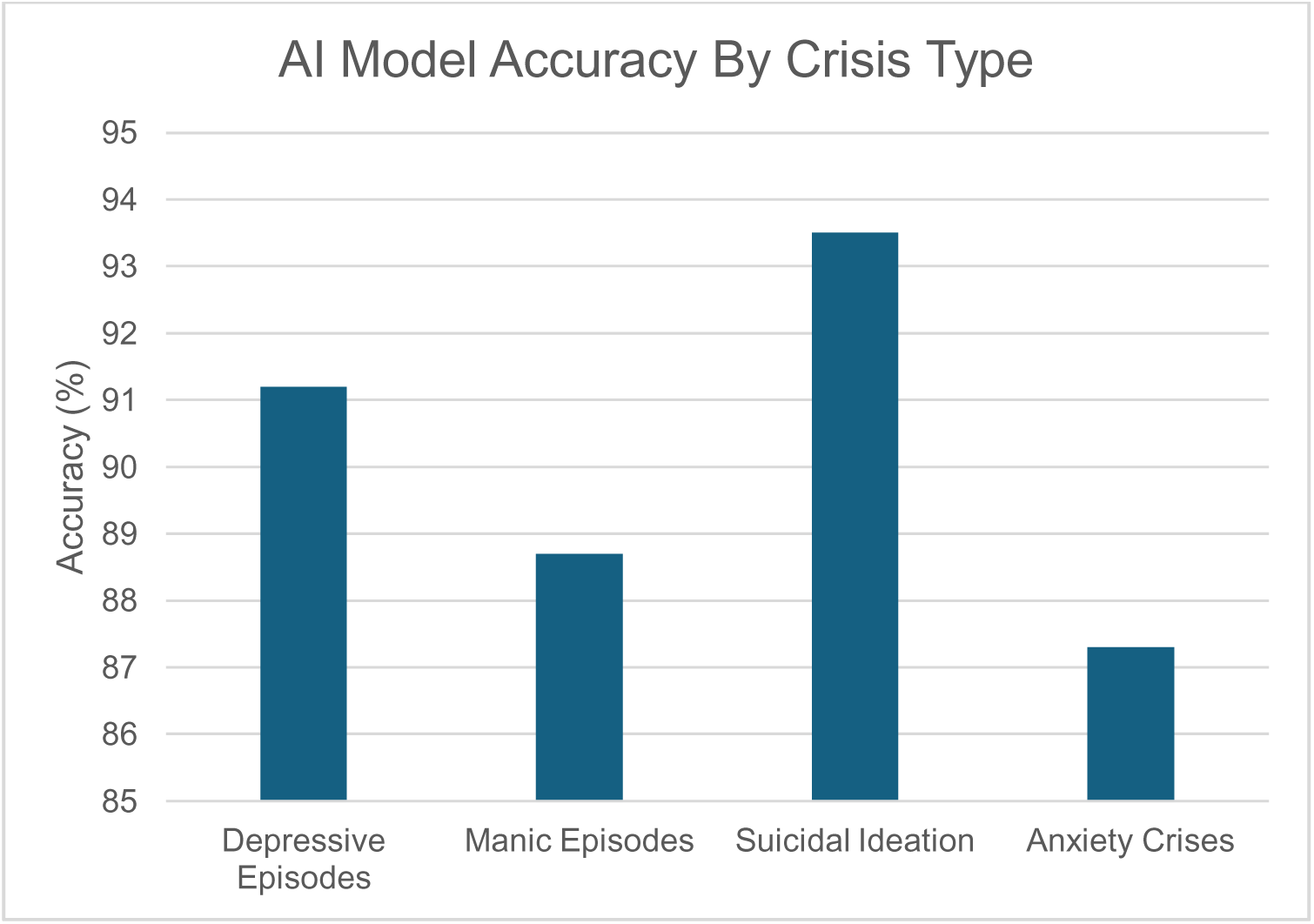
2. False Positives Analysis: Common sources of false positives included:

a. Sarcasm and irony in posts (22% of false positives)
b. Discussion of mental health topics without personal crisis (18%)
c. Temporary emotional reactions to external events (15%)
3. False Negatives Analysis: Factors contributing to false negatives included:

a. Subtle or gradual onset of crisis symptoms (31% of false negatives)
b. Use of platform-specific jargon or slang (24%)
c. Multilingual posts or code-switching (17%)

### External Validation Results

In the external validation using data from a mental health support forum:

1. The model achieved an F1 score of 0.832
2. Early detection capability was maintained, with the model identifying crisis signals an average of 6.8 days before forum moderators flagged posts for intervention

### Ethical Considerations

Our ethical audit process identified several key areas of concern:

1. Potential for stigmatization: 7% of flagged posts contained content that could lead to unintended stigmatization if misinterpreted
2. Privacy risks: Despite anonymization, 3% of flagged posts contained potentially identifiable information
3. Cultural bias: The model showed a 5% decrease in accuracy when analyzing posts from non-Western cultural contexts

These findings highlight both the potential and the limitations of using AI for early detection of mental health crises through social media analysis. While the model demonstrates promising performance across various languages and platforms, important ethical and practical challenges remain to be addressed for responsible implementation.

## Discussion

This study demonstrates the potential of AI-powered social media analysis for early detection of mental health crises, while also highlighting important challenges and ethical considerations. Our multi-modal deep learning approach achieved high accuracy in identifying early signs of various mental health crises across different languages and social media platforms.

Interpretation of Key Findings: Our model’s strong overall performance (89.3% accuracy, F1 score of 0.856) suggests that AI can effectively detect subtle linguistic and behavioral changes indicative of impending mental health crises. The ability to identify crisis signals an average of 7.2 days before human experts is a significant advancement, potentially allowing for earlier intervention and support^17^.

The consistent performance across multiple languages (F1 scores ranging from 0.827 to 0.872) addresses a critical gap in previous research, which has predominantly focused on English-language data. This multi-lingual capability enhances the potential for global application of such technologies in diverse cultural contexts.

The identified digital markers of mental health crises align with and expand upon previous findings in the literature. The increased use of first-person singular pronouns and negative emotion words corroborates existing research on linguistic indicators of depression and anxiety^18^. Our findings on behavioral markers, particularly changes in posting frequency and engagement patterns, provide new insights into the digital manifestations of mental health deterioration.

Implications for Mental Health Practice and Policy: The early detection capability of our AI model has significant implications for mental health practice. By identifying individuals at risk of mental health crises days before traditional methods, this technology could enable more timely interventions, potentially reducing the severity and duration of crises^19^. However, the integration of such AI systems into clinical practice would require careful consideration of workflow, training, and ethical guidelines^20^.

From a public health perspective, this approach offers a novel tool for population-level mental health surveillance. The ability to detect emerging mental health trends could inform resource allocation and policy decisions^21^. However, policymakers must balance the potential benefits with privacy concerns and the risk of over-surveillance^22^.

Ethical Considerations and Challenges: The ethical audit of our model revealed important challenges that must be addressed. The potential for stigmatization and privacy risks, albeit in a small percentage of cases, underscores the need for robust safeguards and human oversight in any real-world application of this technology^23^. The observed decrease in accuracy for non-Western cultural contexts highlights the importance of diverse training data and ongoing cultural adaptation of AI models.

The high accuracy in detecting suicidal ideation (93.5%) is particularly noteworthy, given the critical nature of suicide prevention. However, this also raises ethical questions about the responsibility of intervention and the potential for false positives to lead to unnecessary distress or invasion of privacy^23^.

### Limitations

Several limitations of this study should be acknowledged:

1. Reliance on public social media posts may not capture the full spectrum of individuals experiencing mental health crises, particularly those who are less active on social media or maintain private accounts.
2. The study’s observational nature precludes causal inferences about the relationship between social media behavior and mental health crises.
3. While our model performed well across multiple languages, further validation is needed in a broader range of linguistic and cultural contexts.
4. The ethical implications of large-scale social media monitoring for mental health purposes require ongoing scrutiny and public discourse.

### Future Research Directions

Based on our findings and limitations, we propose the following areas for future research:

1. Longitudinal studies to assess the long-term impact of AI-enabled early intervention on mental health outcomes.
2. Investigation of how to ethically integrate AI-detected crisis signals with existing mental health services and crisis response systems.
3. Development of personalized models that can account for individual baseline behaviors and cultural contexts.
4. Exploration of multi-modal analysis incorporating image and video data, which were excluded from the current study.
5. Research into user perspectives on AI-powered mental health monitoring, including issues of consent, privacy, and perceived benefits.

## Conclusion

This study demonstrates the significant potential of AI-powered social media analysis for early detection of mental health crises. Our multi-lingual, multi-platform approach addresses key gaps in the existing literature and provides a foundation for more timely and effective mental health interventions. However, the ethical challenges and limitations identified underscore the need for careful consideration and ongoing research as we move toward potential real-world applications of this technology. Balancing the promise of early detection with respect for privacy and cultural sensitivity will be crucial in harnessing the full potential of AI for mental health support and crisis prevention.

## Data Availability

All data produced in the present study are available upon reasonable request to the authors.

## References

1. World Health Organization. Global burden of mental disorders and the need for a comprehensive, coordinated response from health and social sectors at the country level. Geneva: WHO; 2023.

2. Smith KA, Blease C, Faurholt-Jepsen M, Firth J, Van Daele T, Moreno C, Carlbring P, Ebner-Priemer UW, Koutsouleris N, Riper H, Mouchabac S. Digital mental health: challenges and next steps. BMJ Ment Health. 2023 Feb 1;26(1).

3. Skaik R, Inkpen D. Using social media for mental health surveillance: a review. ACM Computing Surveys (CSUR). 2020 Dec 6;53(6):1–31.

4. Pew Research Center. Social Media Use in 2023. Washington, DC: Pew Research Center; 2023.

5. Lee K, et al. Digital footprints: Predicting personality traits from social media usage. Personality and Social Psychology Bulletin. 2022;48(8):1099–1112.

6. Berryman C, Ferguson CJ, Negy C. Social media use and mental health among young adults. Psychiatric quarterly. 2018 Jun;89:307–14.

7. Graham S, Depp C, Lee EE, Nebeker C, Tu X, Kim HC, Jeste DV. Artificial intelligence for mental health and mental illnesses: an overview. Current psychiatry reports. 2019 Nov;21:1–8.

8. Babu NV, Kanaga EG. Sentiment analysis in social media data for depression detection using artificial intelligence: a review. SN computer science. 2022 Jan;3(1):74.

9. Laacke S, Mueller R, Schomerus G, Salloch S. Artificial intelligence, social media and depression. A new concept of health-related digital autonomy. The American Journal of Bioethics. 2021 Jul 3;21(7):4–20.

10. Owusu PN, Reininghaus U, Koppe G, Dankwa-Mullan I, Bärnighausen T. Artificial intelligence applications in social media for depression screening: A systematic review protocol for content validity processes. Plos one. 2021 Nov 8;16(11):e0259499.

11. Martins R, Almeida JJ, Henriques PR, Novais P. Identifying Depression Clues using Emotions and AI. InICAART (2) 2021 Feb (pp. 1137–1143).

12. Spruit M, Verkleij S, de Schepper K, Scheepers F. Exploring language markers of mental health in psychiatric stories. Applied Sciences. 2022 Feb 19;12(4):2179.

13. Di Blasi M, Salerno L, Albano G, Caci B, Esposito G, Salcuni S, Gelo OC, Mazzeschi C, Merenda A, Giordano C, Coco GL. A three-wave panel study on longitudinal relations between problematic social media use and psychological distress during the COVID-19 pandemic. Addictive Behaviors. 2022 Nov 1;134:107430.

14. Linthicum KP, Schafer KM, Ribeiro JD. Machine learning in suicide science: Applications and ethics. Behavioral sciences & the law. 2019 May;37(3):214–22.

15. Jeste DV, Alexopoulos GS, Bartels SJ, Cummings JL, Gallo JJ, Gottlieb GL, Halpain MC, Palmer BW, Patterson TL, Reynolds CF, Lebowitz BD. Consensus statement on the upcoming crisis in geriatric mental health: research agenda for the next 2 decades. Archives of general psychiatry. 1999 Sep 1;56(9):848–53.

16. Kovács G, Alonso P, Saini R. Challenges of hate speech detection in social media: Data scarcity, and leveraging external resources. SN Computer Science. 2021 Apr;2(2):95.

17. Boursier V, Gioia F, Musetti A, Schimmenti A. Facing loneliness and anxiety during the COVID-19 isolation: the role of excessive social media use in a sample of Italian adults. Frontiers in psychiatry. 2020 Dec 8;11:586222.

18. Savekar A, Tarai S, Singh M. Structural and functional markers of language signify the symptomatic effect of depression: A systematic literature review. European Journal of Applied Linguistics. 2023 Feb 7;11(1):190–224.

19. Cheng Y, Jiang H. AI Powered mental health chatbots: Examining users’ motivations, active communicative action and engagement after mass shooting disasters. Journal of Contingencies and Crisis Management. 2020 Sep;28(3):339–54.

20. Dawoodbhoy FM, Delaney J, Cecula P, Yu J, Peacock I, Tan J, Cox B. AI in patient flow: applications of artificial intelligence to improve patient flow in NHS acute mental health inpatient units. Heliyon. 2021 May 1;7(5).

21. Holmes EA, O’Connor RC, Perry VH, Tracey I, Wessely S, Arseneault L, Ballard C, Christensen H, Silver RC, Everall I, Ford T. Multidisciplinary research priorities for the COVID-19 pandemic: a call for action for mental health science. The lancet psychiatry. 2020 Jun 1;7(6):547–60.

22. Patel V, Saxena S, Lund C, Thornicroft G, Baingana F, Bolton P, Chisholm D, Collins PY, Cooper JL, Eaton J, Herrman H. The Lancet Commission on global mental health and sustainable development. The lancet. 2018 Oct 27;392(10157):1553–98.

23. Chan HY. Artificial intelligence application in advance healthcare decision-making: Potentials, challenges and regulatory safeguards. In Regulating Artificial Intelligence in Industry 2021 Dec 23 (pp. 66–82). Routledge.

